# Acute and post-acute healthcare utilisation after Delta and Omicron BA.1/2, BA.4/5, XBB infection: a cohort study

**DOI:** 10.1101/2025.10.14.25338047

**Authors:** Jue Tao Lim, Wee Liang En, Reen Wan Li Ho, An Ting Tay, Ervin Zhi Bin Cheong, Julia Regazzini Spinardi, Bulent Nuri Taysi, Karan Thakkar, Moe H Kyaw, Calvin J Chiew, Benjamin Ong, David Chien Boon Lye, Kelvin Bryan Tan

## Abstract

**Background:** Elevated risk of healthcare utilisation in the post-acute phase following SARS-CoV-2 infection is well-documented. However, the burden of acute and post-acute healthcare utilisation following milder Omicron infection is unknown.

**Methods:** We utilized the Singaporean national SARS-CoV-2 testing registry to construct cohorts first infected with SARS-CoV-2 during periods with community transmission predominated by Delta, Omicron BA.1/2, Omicron BA.4/5 or Omicron XBB contrasted versus contemporaneous test-negative controls. We estimate the excess risks/rates/burdens of all-cause (**1**) hospitalisation **(2)** ICU admission (**3**) emergency-department (ED) utilisation and (**4**) total healthcare costs incurred by SARS-CoV-2-infected individuals in acute (0–30 days) and post-acute phases (31–300 days) following infection. Outcomes were compared across variants, vaccination doses and severity of infection.

**Results:** 1,678,678 test-positives were contrasted against 3,346,910 test negatives from 2021–2023. Risks of all-cause hospitalisation, ED visits and overall healthcare costs incurred were significantly higher amongst SARS-CoV-2-infected individuals compared with test-negatives, in both acute and post-acute phase; while risk of ICU admission was elevated only in the acute phase for SARS-CoV-2-infected individuals. In the acute phase, excess risks of healthcare utilisation were elevated 3.6–21.9 times. In the post-acute phase, excess risks of any healthcare utilisation were elevated 0.18–2.02 times, in SARS-CoV-2-infected individuals. Fully vaccinated (2 doses) or boosted (3+ doses) decreased risks and rates of any ED visits, inpatient/ICU admissions and incurred lower excess healthcare costs in both the acute and post-acute period, versus individuals with 0/1 doses.

**Conclusions:** Healthcare utilisation was elevated up to 300 days post-SARS-CoV-2 infection, when compared against test-negatives; even when milder Omicron infection predominated community transmission.

## Introduction

Post-acute sequelae persisting up to years following resolution of acute SARS-CoV-2 infection has been documented in the literature.^1–3^ Increased healthcare utilisation in the post-acute phase following COVID-19 has been documented across multiple population-based cohort studies, resulting in significantly elevated healthcare burden and costs attributed to COVID-19.^4–8^ In the current era of COVID-19 endemicity, reduced severity of acute illness attributed to milder Omicron infection and availability of booster vaccination may potentially translate into lower overall long-term risk and rates of healthcare utilisation, with boosting shown to attenuate both severity of acute illness,^9^ as well as risk of post-acute sequelae following Omicron infection.^10^

However, most studies that explored the association between healthcare utilisation and SARS-CoV-2 infection were conducted predominantly in the pre-Omicron era,^4–8^ and do not fully consider the impacts of vaccination and/or boosting on subsequent healthcare utilisation and costs, particularly long-term healthcare utilisation following recovery from acute illness. In a systematic review evaluating the impact of vaccination on the economic burden of COVID-19 in the United States during the Omicron era, while vaccinated and boosted patients experienced overall reductions in intensive-care-unit (ICU) admissions, length of stay, and emergency-department (ED)/urgent care clinic encounters during the acute phase of illness, information on healthcare utilisation attributable to long-COVID was lacking and identified as a significant gap in the literature.^11^ Population-based studies in the pre-Omicron era comparing differences in healthcare utilisation between infected and uninfected cohorts found that SARS-CoV-2-infected patients had more health care encounters and outpatient visits than uninfected patients 6-12 months post-infection.^12,13,14^ The severity of acute infection was reported to contribute to increased healthcare use in the 6 months post-infection.^15^ In a study of 202,803 US veterans infected in the pre-Omicron era and matched to uninfected controls, outpatient healthcare utilisation was significantly elevated during the 0-30 day peri-infection period; while these differences attenuated over time, they remained significantly elevated at 184-365 days post-infection.^14^ However, these studies were conducted in the pre-Omicron era, significantly limiting generalisability to the current era of Omicron predominance and COVID-19 endemicity.

To address this gap, we enrolled a population-based cohort of adult Singaporeans over periods of pre-Omicron (Delta) and successive Omicron waves (BA.1/2, BA.4/5, XBB), to estimate the acute peri-infection (0-30 days) and post-acute (31-300 days) risks and burdens of healthcare utilisation and costs in SARS-CoV-2-infected individuals versus test-negative controls. Healthcare utilisation was compared across vaccination doses (unvaccinated, fully-vaccinated, boosted) and according to initial severity of acute infection (non-hospitalized, hospitalized, severe).

## Methods

### Study setting and databases

We utilized the national SARS-CoV-2 testing registry from Singapore, a multi-ethnic Southeast Asian city-state, to construct cohorts of adults aged 18 years and above first infected with SARS-CoV-2 during periods with community transmission predominated by Delta, Omicron BA.1/2, Omicron BA.4/5 or Omicron XBB respectively. The Delta variant was first detected in April 2021 and subsequently predominated community transmission (≥90% of sequenced cases on national surveillance) by September 2021; in December 2021, Omicron BA.1/2 was detected in community transmission and co-circulated with Delta, and by January 2022, Omicron BA.1/2 displaced Delta as the predominant strain.^16^ This was followed by a resurgence of cases driven by Omicron BA.4/5 in June 2022 and subsequent emergence of Omicron XBB from October 2022 onwards.^17^ The national healthcare claims database was used to determine healthcare utilization and costs in SARS-CoV-2-infected individuals versus test-negative controls. Vaccination status was obtained from the national vaccination database.

### Exposures and comparators

The primary exposure considered was being test-positive for COVID-19. SARS-CoV-2 infection status (either positive polymerase chain reaction [PCR] or rapid antigen test [RAT]) was determined based on data collected from the national testing registry maintained by the local Ministry of Health (MOH). Throughout the study period, all Singaporeans with symptoms of acute-respiratory-illness (ARI) were strongly encouraged to seek confirmatory testing and clinical assessment at any healthcare provider.^16^ Free SARS-CoV-2 testing was provided at all primary care clinics, including public primary care clinics (polyclinics) and Public Health Preparedness Clinics (PHPCs), which form a nationwide network of more than 1000 private general practitioner (GP) clinics activated to provide subsidized consultation and testing during the pandemic.^16^ Testing for SARS-CoV-2 was mandatory for all individuals who presented with ARI symptoms to any healthcare provider, and positive cases were required by law to be notified to MOH.^15^ Date of notification was taken as T_0._

### Outcomes

The outcomes of interest were total excess acute and post-acute healthcare utilisation in the 30 days following T_0_ and 31 to 300 days following T_0_ respectively. Total healthcare utilisation was broken down as (**1**) all-cause hospitalisation – the length of stay in inpatient settings or the number of unique hospitalisations **(2)** all-cause ICU utilisation – the length of stay in ICU or the number of unique ICU admissions (**3**) all-cause ED utilisation – the total number of unique ED visits. We also examined (**4**) total costs incurred in inpatient settings and (**5**) excess mortality in the respective acute/post-acute periods.

Healthcare utilisation was assessed using the national healthcare claims database. In Singapore, enrolment in the national government administered medical savings scheme (Medisave) is compulsory; Medisave can be claimed against for inpatient care and also outpatient treatment at both public and private healthcare providers.^18^ This enabled comprehensive capture of healthcare utilisation across all care settings (inpatient admissions, ICU admissions, ED visits). Healthcare utilisation in both acute/post-acute phases were compared across vaccination status and severity of initial infection using national databases of COVID-19 vaccination and hospitalisations by subsetting the analysis to that specific subgroup. All healthcare facilities were legally mandated to notify hospitalisations/severe COVID-19 infections.^17,19^ Severe COVID-19 was defined as need for oxygen supplementation or intensive care unit admission.^17,19^ All vaccine doses were recorded in the national vaccination database. BNT162b2(Pfizer) and mRNA-1273(Moderna) were originally approved for use in a two-dose primary series.^19^ Booster vaccinations were rolled out in September 2021; by end November 2021 and 7 March 2022, 24% and 69% had received boosters respectively.^19^

### Study Population and Cohort

A flowchart of cohort construction is provided in **Figure 1**. Individuals who had missing sociodemographic data were excluded. The study population was stratified by period of infection to examine the variant-specific effect of infection on outcomes. Test positives were taken from 1 September – 30 November 2021 (for Delta) and 1 December 2021-31 December 2022 for Omicron. The Omicron-predominant period was stratified into periods of BA.1/2 (1 December 2021-30 March 2022), BA.4/5 (1 June 2022-30 September 2022) and XBB-predominant transmission (18 October 2022-20 Dec 2022). SARS-CoV-2 test-positive individuals were contemporaneously compared against SARS-CoV-2 test-negative individuals (unexposed) drawn from the same population in the respective timeframes with no documented infections prior to the test negative period. Test-negative individuals were assigned an index date T_0_ following the test date. To examine healthcare utilisation in the acute phase, test-positive/negative individuals were followed from T_0_ to T_0_+30 days, and for post-acute healthcare utilisation, from T_0_+31 to T_0_+300 days. For post-acute outcomes, we excluded individuals who died within the first 30 days of the index date, with T_0_ (taken as date of first positive PCR/RAT). We also excluded test-negatives with those had their first SARS-CoV-2 infection within 30 and 300 days of T_0_.

### Covariates

We accounted for differences in baseline characteristics by incorporating the following covariates: demographics (age, sex, ethnicity), number of vaccination doses (0/1,2,3+), comorbidity burden (Charlson’s comorbidity index), socioeconomic status (SES), and ED attendance and inpatient hospitalisations in the preceding 5 years. SES was classified by housing type, a key marker of SES in Singapore.^20^

### Statistical Analysis

Baseline sociodemographic characteristics of the test-positives and respective test-negatives, along with standardized-mean-differences (SMDs) between groups were computed for each period where different variants circulated. We implemented two-part models to ascertain healthcare utilisation and healthcare costs for each study cohort where different variants predominated transmission. In each two-part model, we accounted for imbalances in sociodemographic/comorbidity status in exposed/unexposed groups by propensity score weighting. To do so, logistic regressions were trained taking the outcome variable as COVID-19 test-status, and explanatory variables as vaccination status, age, ethnicity, sex, socioeconomic status, previous healthcare utilisation and comorbidity status. We then used the estimated propensity scores to obtain stabilised inverse probability weights, which were defined as (proportion of test-positive)/propensity scores and (proportion of test-negative)/(1-propensity scores). The doubly-robust approach was used in the two-part models, where the same explanatory variables used to estimate propensity scores were additionally adjusted for in the outcome regressions, to prevent model misspecification in either the propensity score models or the outcome models.

For the two-part model, the first part of the model takes a binomial link function to estimate the probability of any ED visit, inpatient hospitalisation, ICU admission or hospitalisation costs incurred in the follow-up period (T_0_ to T_0_+30, or T_0_+31 to T_0_+300). This enabled us to estimate excess risks in terms of the odds ratios (OR), and quantifies the increase/decrease in acute/post-acute risk of hospital utilisation/incurring of hospital costs given COVID-19 test-positive status. In the second part of the model, we used generalized linear models with the Gamma link function to estimate the (**1**) excess length of stay or number of unique visits in COVID-19 test positives, conditioned on individuals with non-zero inpatient admissions (**2**) excess length of stay or number of unique visits in COVID-19 test positives, conditioned on individuals with non-zero ICU admissions (**3**) number of unique ED visits in COVID-19 test positives, conditioned on individuals with non-zero ED admissions (**4**) excess costs in COVID-19 test positives, conditioned on individuals with non-zero healthcare costs.

For (**1**) – (**3**) we expressed the excess utilisation of unique visits/length of stay in COVID-19 test-positives as a rate ratio, which we defined as the ratio of estimated number of unique visits, length of stay (days) in COVID-19 test-positives over test-negatives among whom had any ED/inpatient/ICU utilisation. Rates were estimated using the second part of the respective two part model. We also expressed excess utilisation of unique visits/length of stay in COVID-19 test-positives in terms of excess burdens, which we defined as the estimated difference in utilisation between COVID-19 test-positives versus test-negatives among whom had any ED/inpatient/ICU utilisation. For (**4**) we similarly defined excess hospital costs in COVID-19 test-positives among as a rate ratio, which we defined as the estimated ratio between the amount of hospital costs incurred in COVID-19 test-positives over test-negatives among those whom incurred any hospital costs. We also expressed excess healthcare costs in COVID-19 test-positives in terms of excess burdens, which we defined as the estimated difference in healthcare costs between COVID-19 test-positives versus test-negatives among whom incurred any healthcare costs. The observed follow-up time was included as offsets in both components of the two-part model.

To examine effect modification, we conducted analyses in subgroups by age (18–64, 65+), ethnicity (Chinese, Malay, Indian or others), sex (male or female), socioeconomic status, vaccination status (0/1, 2, 3+) and disease severity in acute phase of infection (mild, severe).

## Results

### Study population

In total, 1,628,649 adults with new-onset SARS-CoV-2 infection were compared against 3,346,910 test-negatives over successive COVID-19 waves from 2021-2023. 119,170 individuals with new-onset infection during Delta-predominant transmision were compared against 767,017 test-negative individuals; while 833,815 individuals with new-onset infection during Omicron BA.1/2-predominant transmission were compared against 1,072,264 test-negative individuals. During Omicron BA.4/5-predominant transmission, 489,776 individuals had new-onset infection and were compared against 884,516 test-negatives. Finally, during Omicron XBB-predominant transmission, 185,888 individuals had new-onset infection and were compared against 623,113 test-negatives (**Figure 1).**

Baseline sociodemographic and clinical characteristics of infected cases and test-negatived, before and after propensity-score matching, are presented in **Supplementary Tables 1–8**. After weighting, differences in demographic characteristics, socioeconomic status and comorbidity burden between the two groups were small, with all SMDs <0.1 **(Supplementary Tables 1-8).**

### Acute healthcare utilisation and costs

SARS-CoV-2 infected individuals had higher risks of overall healthcare utilisation (any hospitalisations, ED-visits, ICU admissions, and any overall healthcare costs) versus uninfected test-negative controls in the acute peri-infection period spanning 0-30 days from T_0_ (Table 1). Elevated risks of overall healthcare utilisation persisted across pre-Omicron (Delta) periods and throughout subsequent waves of community transmission driven by succeeding Omicron variants (BA.1/2, BA.4/5, XBB). Amongst both test-positive and negative individuals who had any healthcare utilisation, SARS-CoV-2-infected cases had 38.3%–359.8% more ED visits, 51.6%–484.1% more unique inpatient admissions and 35.7%–428.6% more ICU admission, 200.8%–829.0% higher length of stay in inpatient settings and 37.7%–823.2% higher length of stay in ICU. Among those whom incurred hospitalisation costs, SARS-CoV-2-infected individuals had 49.4%–374.9% higher higher hospitalisation costs, which translated to over S$138.1 (95% CI: S$121.9–S$154.3) to S$1047.0 (95% CI: S$980.1–S$1113.8) higher costs per-infected-individual versus test-negatives. In general, risk of overall healthcare utilisation or incurrence of hospitalisation costs in the acute peri-infection period were still elevated in SARS-CoV-2-infected individuals during the Omicron era, but attenuated compared to the pre-Omicron period (Delta). Lower risk of acute healthcare utilisation was also observed amongst individuals infected in subsequent Omicron variant waves (XBB), compared with test-negatives.

**Table 1:**
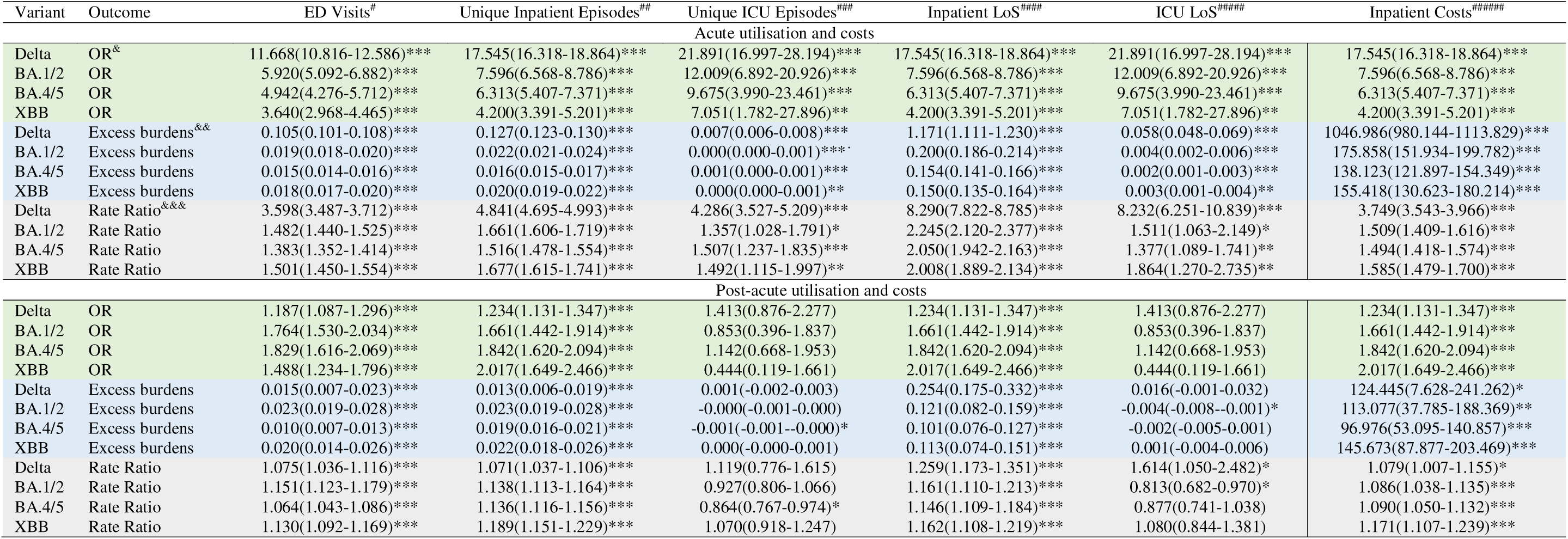
Excess risk and rates of hospital utilisation and costs among test-positive COVID-19 patients versus test-negative controls across different COVID-19 variants/subvariants in the acute and post-acute period. Numbers in parentheses refer to 95% confidence intervals. ^&^Odds ratio estimated from the first part of the two part model. An odds ratio > 1 represents increased risk of any ED visits/inpatient admissions/ICU admissions or incurring of any hospital costs. ^&&^Excess number of ^#^emergency department visits among test-positives versus test-negatives whom had any ED visits ^&&^Excess number of ^##^unique inpatient episodes and ^#####^inpatient length of stay among test-positives versus test-negatives whom had any inpatient admissions ^&&^Excess number of ^###^ICU inpatient episodes and ^######^ICU length of stay among test-positives versus test-negatives whom had any inpatient admissions ^&&^Excess costs of among ^#####^test-positives versus test-negatives whom had any hospital costs incurred ^&&^Any value >0 represents higher utilisation levels among COVID-19 test-positives versus test-negatives ^&&&^Excess proportion of ^#^emergency department visits among test-positives versus test-negatives whom had any ED visits ^&&&^Excess proportion of ^##^unique inpatient episodes and ^#####^inpatient length of stay among test-positives versus test-negatives whom had any inpatient admissions ^&&&^Excess proportion of ^###^ICU inpatient episodes and ^######^ICU length of stay among test-positives versus test-negatives whom had any inpatient admissions ^&&&^Excess proportion of costs of among ^#####^test-positives versus test-negatives whom had any hospital costs incurred ^&&&^Any value >1 represents higher utilisation levels among COVID-19 test-positives versus test-negatives. (1-Rate Ratio)*100 is the percentage increase in utilisation in the test-positive group. Statistical significance at *0.05, **0.01,***0.001 level.

### Post-acute healthcare utilisation and cost

In the post-acute period spanning 31-300 days from T_0_, SARS-CoV-2 infected individuals had 18.7%–82.9% higher risk of ED visits, 23.4–201.7% higher risk of inpatient admissions and 23.4%–201.7% higher risk of incurring any costs versus test-negative controls. During the post-acute period, higher risks of overall healthcare utilisation (any hospitalisations, ED-visits, ICU admissions, and any overall healthcare costs) were observed in SARS-CoV-2-infected individuals compared with test-negatives, with the exception of ICU admissions (Table 1). Risks of all-cause hospitalisations and incurrence of any hospitalisation cost amongst SARS-CoV-2-infected individuals, versus test-negatives, in the post-acute period were numerically higher during transmission predominated by successive Omicron variants, versus the pre-Omicron period (Delta). When we compared test-positives versus negatives who had any hospitalisation in the post-acute period, those infected during Delta had higher length-of-stay but lower number of unique hospitalisations.(Table 1)

### Acute healthcare utilisation and cost by vaccination status

Vaccination and boosting drastically decreased the risk and level of hospital utilisation among COVID-19 test positives in the acute period of infection across both Delta, Omicron BA.1/2, BA.4/5 and XBB. Comparing individuals with 0/1 vaccination doses versus those who had 2 or 3+ vaccination doses, we found that test-positives who were fully vaccinated (2 doses) or boosted (3 doses or more), had far lower risk of any ED visits, inpatient/ICU admissions or incurring any costs. Furthermore, among test-positive and negative individuals who had any ED visits, inpatient/ICU admissions or incurred any costs, infectees of any vaccination status had elevated hospital utilisation, but those whom were boosted or fully vaccinated had lower excess burdens/rates of ED visits, unique inpatient/ICU episodes, lowered length of stay in inpatient or ICU settings and lowered healthcare costs incurred, versus individuals with no/partial vaccination. These findings persisted across Delta, Omicron BA.1/2, BA.4/5 and XBB (Table 2). Excess healthcare costs in the acute phase among test-positives whom had any healthcare utilisation were approximately reduced 5-fold or more for fully vaccinated individuals and reduced 10-fold or more for boosted individuals, versus unvaccinated test-positives.

**Table 2:** Excess risk and rates of hospital utilisation and costs among test-positive COVID-19 patients versus test-negative controls across different COVID-19 variants/subvariants in the acute period by vaccination doses. Numbers in parentheses refer to 95% confidence intervals. ^&^Odds ratio estimated from the first part of the two part model. An odds ratio > 1 represents increased risk of any ED visits/inpatient admissions/ICU admissions or incurring of any hospital costs. ^&&^Excess number of ^#^emergency department visits among test-positives versus test-negatives whom had any ED visits ^&&^Excess number of ^##^unique inpatient episodes and ^#####^inpatient length of stay among test-positives versus test-negatives whom had any inpatient admissions ^&&^Excess number of ^###^ICU inpatient episodes and ^######^ICU length of stay among test-positives versus test-negatives whom had any inpatient admissions ^&&^Excess costs of among ^#####^test-positives versus test-negatives whom had any hospital costs incurred ^&&^Any value >0 represents higher utilisation levels among COVID-19 test-positives versus test-negatives ^&&&^Excess proportion of ^#^emergency department visits among test-positives versus test-negatives whom had any ED visits ^&&&^Excess proportion of ^##^unique inpatient episodes and ^#####^inpatient length of stay among test-positives versus test-negatives whom had any inpatient admissions ^&&&^Excess proportion of ^###^ICU inpatient episodes and ^######^ICU length of stay among test-positives versus test-negatives whom had any inpatient admissions ^&&&^Excess proportion of costs of among ^#####^test-positives versus test-negatives whom had any hospital costs incurred ^&&&^Any value >1 represents higher utilisation levels among COVID-19 test-positives versus test-negatives. (1-Rate Ratio)*100 is the percentage increase in utilisation in the test-positive group. Statistical significance at *0.05, **0.01,***0.001 level.

| Acute utilisation and costs (T<sub>0</sub> to T<sub>0</sub>+30) |  |  |  |  |  |  |  |
| --- | --- | --- | --- | --- | --- | --- | --- |
| Variant | Outcome | ED Visits<sup>#</sup> | Unique Inpatient Episodes<sup>##</sup> | Unique ICU Episodes<sup>###</sup> | Inpatient<sup>####</sup> | ICU<sup>#####</sup> | Inpatient Costs<sup>#####</sup> |
| Vaccination Dose = 0/1 |  |  |  |  |  |  |  |
| Delta | OR<sup>&</sup> | 11.791(10.895-12.761)*** | 18.385(16.974-19.913)*** | 21.112(16.524-26.975)*** | 18.385(16.974-19.913)*** | 21.112(16.524-26.975)*** | 18.385(16.974-19.913)*** |
| BA.1/2 | OR | 5.307(4.658-6.046)*** | 6.691(5.872-7.625)*** | 10.060(5.706-17.736)*** | 6.691(5.872-7.625)*** | 10.060(5.706-17.736)*** | 6.691(5.872-7.625)*** |
| BA.4/5 | OR | 4.679(3.994-5.483)*** | 6.370(5.353-7.581)*** | 10.913(4.624-25.757)*** | 6.370(5.353-7.581)*** | 10.913(4.624-25.757)*** | 6.370(5.353-7.581)*** |
| XBB | OR | 3.472(2.794-4.315)*** | 4.039(3.217-5.071)*** | - | 4.039(3.217-5.071)*** | - | 4.039(3.217-5.071)*** |
| Delta | Excess burdens<sup>&&</sup> | 0.368(0.348-0.387)*** | 0.472(0.453-0.491)*** | 0.048(0.040-0.057)*** | 5.422(5.095-5.748)*** | 0.453(0.346-0.559)*** | 6006.849(5469.382-6544.317)*** |
| BA.1/2 | Excess burdens | 0.162(0.146-0.179)*** | 0.201(0.183-0.218)*** | 0.011(0.007-0.015)*** | 2.192(1.833-2.552)*** | 0.062(0.031-0.094)*** | 2366.899(2020.786-2713.013)*** |
| BA.4/5 | Excess burdens | 0.174(0.147-0.201)*** | 0.218(0.188-0.247)*** | 0.034(0.007-0.061)* | 2.105(1.785-2.425)*** | 0.125(0.028-0.222)* | 2478.908(1999.468-2958.347)*** |
| XBB | Excess burdens | 0.144(0.110-0.178)*** | 0.157(0.121-0.192)*** | - | 1.819(1.385-2.254)*** | - | 2190.887(1495.708-2886.066)*** |
| Delta | Rate Ratio<sup>&&&</sup> | 8.407(7.831-9.025)*** | 11.073(10.377-11.816)*** | 22.962(17.471-30.177)*** | 23.234(21.192-25.473)*** | 46.905(28.929-76.053)*** | 14.247(12.699-15.983)*** |
| BA.1/2 | Rate Ratio | 4.928(4.289-5.662)*** | 6.384(5.589-7.293)*** | 11.045(6.123-19.923)*** | 8.233(6.779-10.000)*** | 13.801(6.408-29.722)*** | 7.097(5.898-8.540)*** |
| BA.4/5 | Rate Ratio | 4.712(3.930-5.650)*** | 6.934(5.688-8.453)*** | 64.694(8.827-474.173)*** | 9.248(7.112-12.026)*** | 36.839(4.603-294.860)*** | 8.318(6.316-10.956)*** |
| XBB | Rate Ratio | 3.726(2.959-4.693)*** | 4.266(3.329-5.466)*** | - | 6.436(4.701-8.813)*** | - | 5.920(4.295-8.159)*** |
| Vaccination Dose = 2 |  |  |  |  |  |  |  |
| Delta | OR | 3.720(3.588-3.857)*** | 5.073(4.891-5.262)*** | 3.977(3.271-4.836)*** | 5.073(4.891-5.262)*** | 3.977(3.271-4.836)*** | 5.073(4.891-5.262)*** |
| BA.1/2 | OR | 1.988(1.881-2.102)*** | 2.311(2.177-2.455)*** | 1.818(1.151-2.872)* | 2.311(2.177-2.455)*** | 1.818(1.151-2.872)* | 2.311(2.177-2.455)*** |
| BA.4/5 | OR | 1.922(1.767-2.091)*** | 2.427(2.210-2.666)*** | 2.051(1.221-3.443)** | 2.427(2.210-2.666)*** | 2.051(1.221-3.443)** | 2.427(2.210-2.666)*** |
| XBB | OR | 1.981(1.709-2.297)*** | 2.276(1.938-2.674)*** | 0.657(0.218-1.982) | 2.276(1.938-2.674)*** | 0.657(0.218-1.982) | 2.276(1.938-2.674)*** |
| Delta | Excess burdens | 0.081(0.077-0.084)*** | 0.094(0.090-0.097)*** | 0.003(0.002-0.003)*** | 0.686(0.643-0.729)*** | 0.021(0.014-0.027)*** | 494.589(454.604-534.573)*** |
| BA.1/2 | Excess burdens | 0.038(0.034-0.042)*** | 0.047(0.043-0.051)*** | 0.001(0.000-0.001)* | 0.339(0.306-0.372)*** | 0.003(0.000-0.005)* | 270.120(212.838-327.402)*** |
| BA.4/5 | Excess burdens | 0.047(0.040-0.054)*** | 0.061(0.053-0.069)*** | 0.002(0.001-0.004)** | 0.520(0.427-0.613)*** | 0.011(0.003-0.018)** | 577.592(475.446-679.739)*** |
| XBB | Excess burdens | 0.045(0.033-0.057)*** | 0.055(0.043-0.068)*** | 0.000(-0.002-0.003) | 0.517(0.363-0.670)*** | 0.007(-0.010-0.023) | 414.316(252.169-576.463)*** |
| Delta | Rate Ratio | 3.412(3.291-3.536)*** | 4.627(4.465-4.795)*** | 3.989(3.184-4.997)*** | 7.522(7.096-7.974)*** | 8.158(5.923-11.237)*** | 3.377(3.163-3.606)*** |
| BA.1/2 | Rate Ratio | 1.974(1.860-2.094)*** | 2.369(2.222-2.526)*** | 1.767(1.095-2.852)* | 2.975(2.737-3.234)*** | 2.109(1.199-3.710)** | 1.800(1.621-1.999)*** |
| BA.4/5 | Rate Ratio | 1.968(1.791-2.163)*** | 2.678(2.415-2.970)*** | 2.764(1.574-4.853)*** | 3.056(2.579-3.622)*** | 3.746(1.770-7.928)*** | 2.840(2.432-3.317)*** |
| XBB | Rate Ratio | 1.925(1.645-2.254)*** | 2.389(2.008-2.843)*** | 1.083(0.357-3.291) | 3.092(2.385-4.007)*** | 1.649(0.545-4.989) | 2.113(1.652-2.701)*** |
| Vaccination Dose = 3+ |  |  |  |  |  |  |  |
| Delta | OR | 2.672(2.468-2.892)*** | 3.430(3.189-3.691)*** | 2.146(1.430-3.221)*** | 3.430(3.189-3.691)*** | 2.146(1.430-3.221)*** | 3.430(3.189-3.691)*** |
| BA.1/2 | OR | 1.379(1.327-1.433)*** | 1.516(1.448-1.588)*** | 1.221(0.865-1.723) | 1.516(1.448-1.588)*** | 1.221(0.865-1.723) | 1.516(1.448-1.588)*** |
| BA.4/5 | OR | 1.333(1.305-1.362)*** | 1.355(1.324-1.388)*** | 1.245(1.071-1.448)** | 1.355(1.324-1.388)*** | 1.245(1.071-1.448)** | 1.355(1.324-1.388)*** |
| XBB | OR | 1.458(1.412-1.505)*** | 1.557(1.505-1.612)*** | 1.437(1.136-1.818)** | 1.557(1.505-1.612)*** | 1.437(1.136-1.818)** | 1.557(1.505-1.612)*** |
| Delta | Excess burdens | 0.058(0.051-0.065)*** | 0.081(0.074-0.089)*** | 0.002(0.000-0.003)* | 0.917(0.649-1.186)*** | 0.015(0.004-0.026)** | 517.699(416.347-619.051)*** |
| BA.1/2 | Excess burdens | 0.012(0.010-0.013)*** | 0.015(0.013-0.016)*** | 0.000(-0.000-0.000) | 0.126(0.112-0.141)*** | 0.001(-0.000-0.002) | 95.566(70.458-120.673)*** |
| BA.4/5 | Excess burdens | 0.012(0.011-0.013)*** | 0.012(0.011-0.013)*** | 0.000(0.000-0.000)** | 0.117(0.106-0.128)*** | 0.001(-0.000-0.002) | 90.583(75.817-105.349)*** |
| XBB | Excess burdens | 0.016(0.014-0.018)*** | 0.017(0.016-0.019)*** | 0.000(0.000-0.001)* | 0.121(0.108-0.135)*** | 0.002(0.001-0.004)** | 125.842(102.458-149.225)*** |
| Delta | Rate Ratio | 2.538(2.337-2.756)*** | 3.239(3.007-3.489)*** | 2.196(1.417-3.404)*** | 6.888(5.261-9.018)*** | 4.385(2.352-8.175)*** | 2.751(2.408-3.144)*** |
| BA.1/2 | Rate Ratio | 1.370(1.317-1.424)*** | 1.547(1.476-1.620)*** | 1.295(0.897-1.870) | 2.113(1.967-2.270)*** | 1.347(0.866-2.094) | 1.443(1.323-1.573)*** |
| BA.4/5 | Rate Ratio | 1.344(1.314-1.375)*** | 1.456(1.419-1.495)*** | 1.385(1.131-1.695)** | 1.985(1.877-2.098)*** | 1.276(0.998-1.631) | 1.429(1.354-1.508)*** |
| XBB | Rate Ratio | 1.475(1.423-1.528)*** | 1.641(1.579-1.706)*** | 1.471(1.088-1.989)* | 1.958(1.839-2.084)*** | 1.882(1.267-2.796)** | 1.554(1.447-1.670)*** |

### Post-acute healthcare utilisation and cost by vaccination status

In the post-acute period, vaccination and boosting drastically decreased risk and level of healthcare utilisation among COVID-19 test positives in Delta, Omicron BA.1/2, BA.4/5 and XBB (Table 3). Comparing individuals with 0/1 vaccination doses versus those who had 2 or 3+ vaccination doses, we found that test-positives who were fully vaccinated (2 doses) or boosted (3 doses or more), had modestly decreased risks of any ED visits, inpatient admissions or incurring any costs in the post-acute period, versus individuals with 0/1 doses. Among test-positive and negative individuals who had any ED visits, inpatient/ICU admissions or incurring any costs, while infectees of any vaccination status had elevated hospital utilisation, those whom were boosted or fully vaccinated had lowered rates of ED visits, unique inpatient/ICU episodes, lowered length of stay in inpatient or ICU settings and lowered healthcare costs incurred versus unvaccinated individuals. In particular, among individuals who incur any healthcare costs, excess healthcare costs in the post-acute phase among test-positives whom had any hospital utilisation were approximately reduced 30% or more for fully vaccinated individuals and halved or more for boosted individuals, versus unvaccinated test-positives.

**Table 3:**
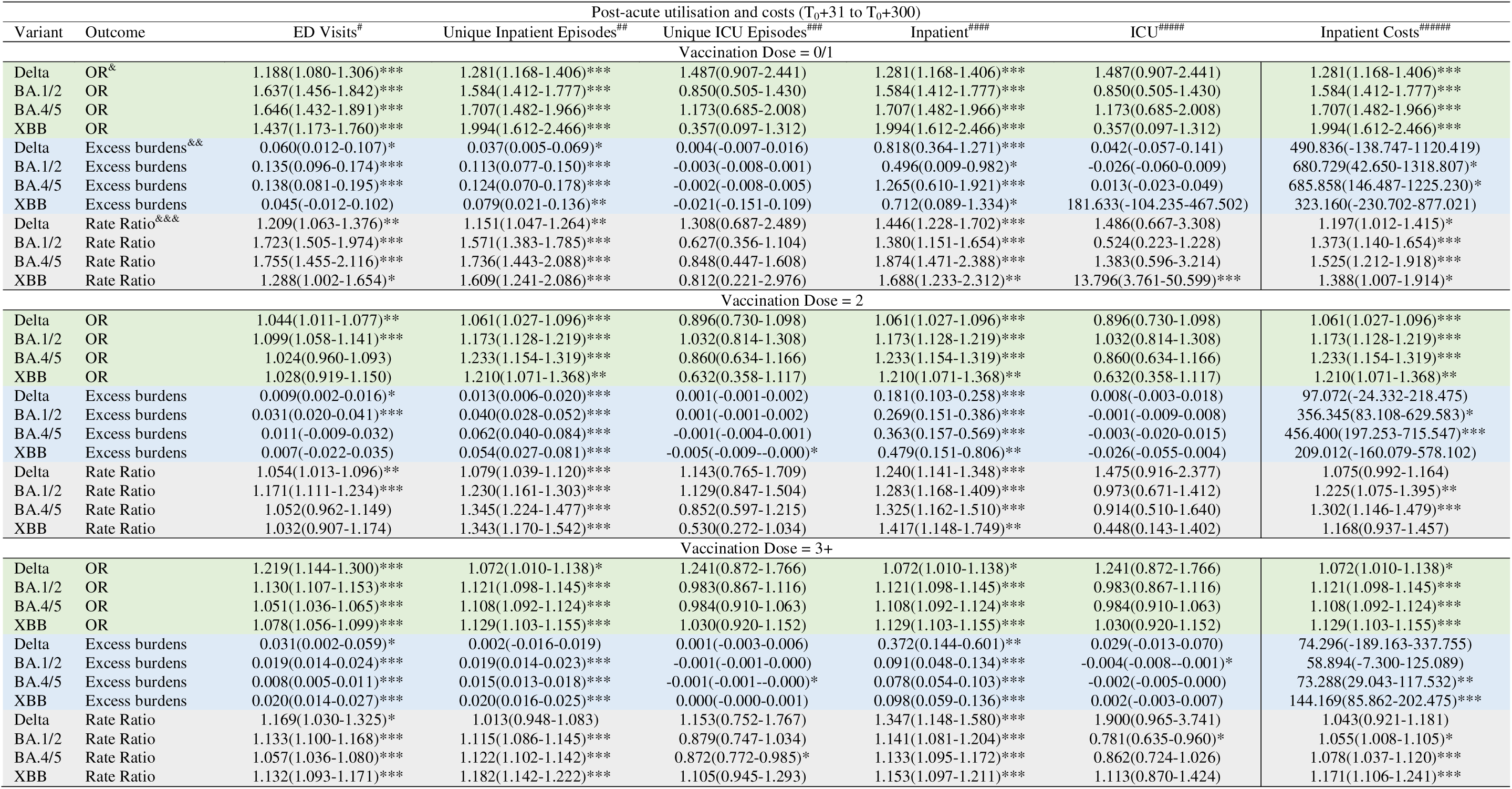
Excess risk and rates of hospital utilisation and costs among test-positive COVID-19 patients versus test-negative controls across different COVID-19 variants/subvariants in the post-acute period by vaccination doses. Numbers in parentheses refer to 95% confidence intervals. ^&^Odds ratio estimated from the first part of the two part model. An odds ratio > 1 represents increased risk of any ED visits/inpatient admissions/ICU admissions or incurring of any hospital costs. ^&&^Excess number of ^#^emergency department visits among test-positives versus test-negatives whom had any ED visits ^&&^Excess costs of among ^#####^test-positives versus test-negatives whom had any hospital costs incurred ^&&^Any value >0 represents higher utilisation levels among COVID-19 test-positives versus test-negatives ^&&&^Excess proportion of ^#^emergency department visits among test-positives versus test-negatives whom had any ED visits ^&&&^Excess proportion of ^##^unique inpatient episodes and ^#####^inpatient length of stay among test-positives versus test-negatives whom had any inpatient admissions ^&&&^Excess proportion of ^###^ICU inpatient episodes and ^######^ICU length of stay among test-positives versus test-negatives whom had any inpatient admissions ^&&&^Excess proportion of costs of among ^#####^test-positives versus test-negatives whom had any hospital costs incurred ^&&&^Any value >1 represents higher utilisation levels among COVID-19 test-positives versus test-negatives. (1-Rate Ratio)*100 is the percentage increase in utilisation in the test-positive group. Statistical significance at *0.05, **0.01,***0.001 level.

We explored potential effect modification of gender, ethnicity, age and SES on SARS-CoV-2 infection and post-acute hospital utilisation by subsetting our analysis to those specific strata and examining excess risks, rates and burdens in each subgroup **(Supplementary Tables 9-21).** In general, gender and ethnicity did not modify estimates of excess risks, rates and burdens of hospital utilisation significantly. However, older individuals aged 65 and above had higher risks of healthcare utilisation in both acute and post-acute periods, when infected with SARS-CoV-2, versus younger adults aged 18–64. This pattern was also found when comparing across comorbidity and socio-economic status subgroups, where test-positives with 1 or more comorbidities had higher excess risks, rates and burdens of healthcare utilisation in both acute/post-acute periods versus test positives with 0 comorbidites. Similarly, individuals who were of higher socio-economic status (i.e living in private property, 4-5 room public apartments), had higher healthcare utilisation versus individuals living in 1-3 room public apartments.

Across the severity gradient of acute-phase infection, we found that acute and post-acute excess risk, rates and burdens of hospital utilisation were primarily driven by individuals who had severe SARS-CoV-2 infection. In the acute period, the risk of having any ED visits, inpatient admissions, ICU admissions or incurring healthcare costs were 10–100 fold larger for severe test-positives versus mild test-positives. Similarly, rates and excess burdens of hospital utilisation among those who had one or more ED visits or inpatient/ICU admissions were larger in severe SARS-CoV-2 test positives versus mild infectees. Across different SARS-CoV-2 variants, we found that the estimates of excess risks of any having post-acute ED visits, inpatient admissions, ICU admissions or incurring healthcare costs were mixed, for mild SARS-CoV-2 test positives versus test-negatives; consequentially, we did not notice a consistent pattern of excess burdens among mild SARS-CoV-2 test positives versus test-negatives across different variants. However, elevated post-acute risks, burdens and rates of hospital utilisation were found for severe SARS-CoV-2 infectees.

## Discussion

In a highly vaccinated and boosted cohort of 1,628,649 adult Singaporeans first infected with SARS-CoV-2 during Delta-predominant transmission and succeeding Omicron waves (BA.1/2, BA.4/5, XBB), we found significantly elevated risks of any emergency department visits, inpatient admissions and incurring any inpatient costs in both the acute and post-acute periods, versus 3,346,910 contemporary, test-negative controls. In the acute peri-infection period (0-30 days), excess risks of any healthcare utilisation were elevated by 3.6–21.9 times, and in the post-acute period (31-300 days), excess risks of any healthcare utilisation were elevated by 0.18–2.02 times versus uninfected test-negatives. Severity of acute infection was a key driver for the elevated risk of healthcare utilization and costs in both peri-acute and post-acute phase, with subgroup analyses demonstrating that the risk of having any ED visits, inpatient admissions, ICU admissions or incurring healthcare costs was 10–100 fold larger for severe COVID-19 infections versus mild cases.

Healthcare utilisation in the acute peri-infection period was substantially lower during Omicron transmission, compared with the pre-Omicron (Delta) period; likely due to milder symptoms and reduced severity of acute infection with successive Omicron variants.^21^ However, despite milder acute infection, estimates of post-acute healthcare utilisation following Omicron infection, versus test-negatives, were numerically higher during transmission predominated by successive Omicron variants, versus the pre-Omicron period (Delta). These findings were consistent with other studies that compared differences in healthcare utilisation between infected and uninfected cohorts in the pre-Omicron era, and found that a diagnosis of COVID-19 was associated with increased healthcare utilization and costs up to six-months post-diagnosis.^13–15^ While the acute burden of COVID-19 on healthcare has dramatically declined with the emergence of Omicron, resulting in substantial decrease in use of acute COVID-19-related inpatient and outpatient care services,^22,23^ post-acute sequelae of COVID-19 can still impose a prolonged burden on healthcare systems in the Omicron era. Healthcare utilisation post-COVID-19 was only modestly increased, in line with prior studies in our local population showing only modest burden of cardiovascular, neuropsychiatric and autoimmune complications up to 300 days following SARS-CoV-2 infection.^24–26^ However, the large number of individuals infected with SARS-CoV-2 during Omicron means that even small increases can still translate into a substantial rise in healthcare costs. Healthcare policy-makers need to anticipate these excess costs and allocate resources accordingly during the transition toward COVID-19 endemicity.

Vaccination and boosting generally decreased both acute and post-acute risks of healthcare utilisation, as well as the excess rates/burdens of healthcare utilisation if these resources were utilized. Crucially, apart from conferring protection against severe COVID-19 disease and hospitalisation attributable to Omicron infection in the acute setting,^19,27^ vaccination and boosting was previously demonstrated to reduce the incidence of prolonged post-COVID-19 sequelae across multiple organ systems, following Omicron infection.^10,24–26^ In our local population, receipt of booster doses was associated with attenuated risk of new-incident cardiovascular, neuropsychiatric and autoimmune complications amongst adults infected during Omicron BA.1/2 predominance.^24–26^ This is likely due to the impact of vaccination and boosting on reducing severity of initial infection;^19^ a graded increase in post-acute healthcare utilisation by severity of initial infection was also noted in prior studies during pre-Omicron pandemic waves.^7–8,13–15^ One recent US study reported a lower incidence of post-acute sequelae in Omicron period compared to the pre-Omicron period, with 70% of this decrease attributed to the impact of vaccination.^27^ Our findings demonstrate the key role of booster vaccinations in public health strategies aimed at mitigating potential excess healthcare utilisation attributable to both acute and post-acute periods,^28^ during COVID-19 endemicity.

Our study has several strengths. We used nationally representative electronic health databases, combined with individually resolved data on testing and vaccination statuses to estimate the post-acute excess risks and rates of hospital utilisation for COVID-19 patients across vaccination status and periods where different variants predominated in SARS-CoV-2 community transmission. We analytical framework without adjustment of covariates in the regression step for analysis, and found that our HRs were replicable.

This study has several limitations: (**1**)the unexposed groups may be contaminated by undiagnosed or asymptomatic infections; while we used comprehensive national testing databases to reduce the likelihood of misclassification, misclassification would bias estimated ORs, IRRs and excess burdens downwards, inducing more conservative estimates. While results of self-administered RAT were not recorded in the national database, individuals with positive results on self-administered RAT were strongly encouraged to proceed to primary care providers for confirmatory PCR/RAT testing and clinical assessment, which were widely available and provided free-of-charge.^16^ (**2**)While our study comprised a nationally representative multi-ethnic Asian population, it is predominantly of Chinese ethnicity, which may limit the generalizability of study findings. (**3**)The epidemiology of post-acute sequelae after SARS-CoV-2 infection may also change, due to the emergence of new variants as well as the subsequent introduction of COVID-19 therapeutics. In addition, the duration of post-acute COVID-19 is unknown and therefore our results may have been under or overestimates.

In conclusion, utilizing data from a multi-ethnic nationwide cohort of adult Singaporeans infected during waves of Delta/Omicron BA.1/2 transmission, we found increased risk and rates of healthcare utilisation in the 300 days post-SARS-CoV-2 infection when compared against contemporary test-negatives. Vaccination and boosting can potentially mitigate the risk of excess healthcare utilisation attributable to post-acute sequelae of COVID-19.

## Data Availability

The databases with individual-level information used for this study are not publicly available due to personal data protection. Deidentified data can be made available for research, subject to approval by the Ministry of Health of Singapore. All inquiries should be sent to the corresponding author.

## Declarations

### Ethics approval and consent to participate

This study was done as part of national public health research under the Infectious Diseases Act, Singapore. Need for consent to participate was waived and deemed unnecessary, and ethics review by an institutional review board was not required under the Infectious Diseases Act, Singapore

### Consent for publication

Not applicable

### Competing interests

Ervin Zhi Bin Cheong, Julia Regazzini Spinardi, Bulent Nuri Taysi, Karan Thakkar, Moe H Kyaw are paid employees of Pfizer. All other authors report no conflicts of interest.

### Funding

LEW is supported by the National Medical Research Council through the Clinician-Scientist New Investigator Grant (2024). JTL is supported by the Ministry of Education (MOE), Singapore Start-up Grant.

### Authors’ contributions

DL, KBT, JS, MHK conceived the study idea and contributed to the study protocol. JTL, LEW contributed to literature search and writing of the manuscript. All authors contributed to critical review and editing of the manuscript. DCL and KBT provided supervision. KBT, LEW, JTL, RWLH, KT, JS, MHK and ATT contributed to study design. ATT and RWLH contributed to data collection, and ATT, JTL and RWLH contributed to data analysis. All authors had full access to all the data in the study and take responsibility for the decision to submit for publication. LEW, JTL, RWLH and ATT directly accessed and verified the underlying data reported in the manuscript.

### Clinical Trial

Not applicable

## Acknowledgements

We thank Florence Lefebvre d’Hellencourt, Carlos Fernando Mendoza, Srinivas Rao Valluri, Caroline White and Egemen Ozbilgili from Pfizer for their support and assistance in this project.

## Notes

### Author Declarations

This study was done as part of national public health research under the Infectious Diseases Act, Singapore. Need for consent to participate was waived and deemed unnecessary, and ethics review by an institutional review board was not required under the Infectious Diseases Act, Singapore. The datasets are de-identified prior to use in study

